# Genetic, structural and clinical analysis of spastic paraplegia 4

**DOI:** 10.1101/2021.07.20.21259482

**Authors:** Parizad Varghaei, Mehrdad A Estiar, Setareh Ashtiani, Simon Veyron, Kheireddin Mufti, Etienne Leveille, Eric Yu, Dan Spiegelman, Marie-France Rioux, Grace Yoon, Mark Tarnopolsky, Kym M. Boycott, Nicolas Dupre, Oksana Suchowersky, Jean-François Trempe, Guy A. Rouleau, Ziv Gan-Or

## Abstract

**Introduction:** Spastic paraplegia type 4 (SPG4), resulting from heterozygous mutations in the *SPAST* gene, is the most common form among the heterogeneous group of hereditary spastic paraplegias (HSPs). We aimed to study genetic and clinical characteristics of SPG4 across Canada.

**Methods:** The *SPAST* gene was analyzed in a total of 696 HSP patients from 431 families by either HSP-gene panel sequencing or whole exome sequencing (WES). We used Multiplex ligation-dependent probe amplification to analyze copy number variations (CNVs), and performed *in silico* structural analysis of selected mutations. Clinical characteristics of patients were assessed, and long-term follow-up was done to study genotype-phenotype correlations.

**Results:** We identified 157 SPG4 patients from 65 families who carried 41 different *SPAST* mutations, six of which are novel and six are CNVs. We report novel aspects of mutations occurring in Arg499, a case with homozygous mutation, a family with probable compound heterozygous mutations, three patients with *de novo* mutations, three cases with pathogenic synonymous mutation, co-occurrence of SPG4 and clinically isolated syndrome, and novel or rarely reported signs and symptoms seen in SPG4 patients.

**Conclusion:** Our study demonstrates that SPG4 is a heterogeneous type of HSP, with diverse genetic features and clinical manifestations. In rare cases, biallelic inheritance, *de novo* mutation, pathogenic synonymous mutations and CNVs should be considered.

## Introduction

Spastic paraplegia type 4 (SPG4, OMIM #182601) is the most frequent form of hereditary spastic paraplegias (HSPs), caused by heterozygous mutations in the *SPAST* gene. With more than 80 potentially causative loci or genes reported to date, HSPs are known to affect about 1.8/100,000 of the population,^1^ and autosomal dominant (AD) HSPs comprise 43%-80% of them.^2, 3^ Among all AD-HSPs, 70-80% are categorized as “pure” with a phenotype limited to pyramidal signs in the lower limbs, with or without deep sensory loss and sphincter disturbances.^1^ Of all pure AD-HSPs, about 40% are caused by *SPAST* mutations.^1^

*SPAST* encodes spastin, which is a protein from the AAA (ATPase associated with various cellular activities) family of ATPases. Spastin controls different aspects of microtubule dynamics (e.g. microtubule number, motility, length, disassembly and remodeling), and hydrolyses ATP to cleave microtubules, a necessary step in axonal transport.^4, 5^ The structure of human spastin residues 323-610 was solved by cryoelectron microscopy revealing an AAA+ ATPase homohexamer (Figure 1A), with ATPase active sites located at the interface between every adjacent subunit (Figure 1B).^5, 6^

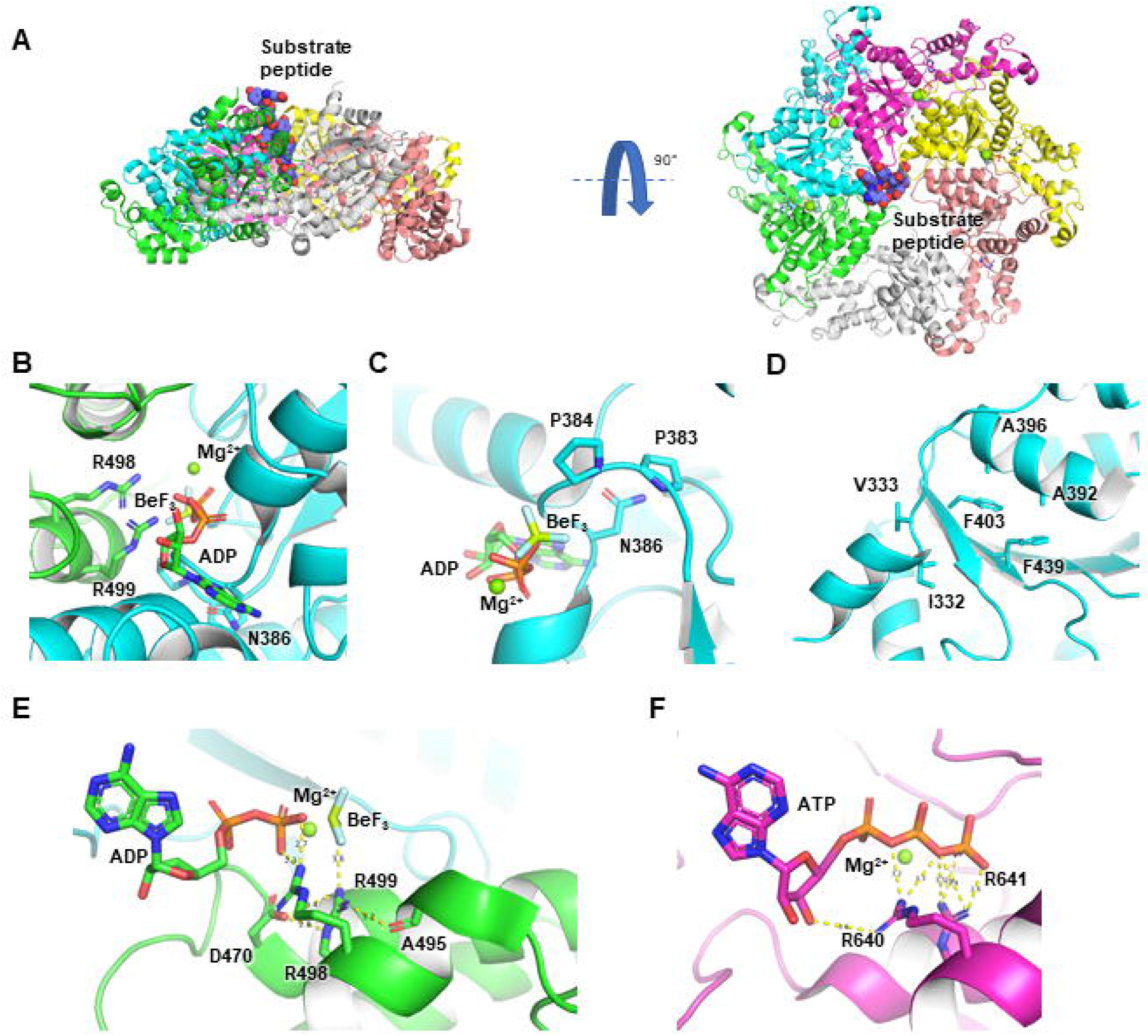

To date, more than 200 *SPAST* mutations have been found.^7^. Exon deletions may account for 20% of cases in whom point mutations are not detected.^8^

The penetrance of SPG4 is up to 80-90% and is age-dependent,^9^ with age at onset (AAO) that may range from infancy to the eighth decade of life. Most cases present as juvenile or adult-onset pure spastic paraplegia with urinary sphincter disturbances, and pes cavus. However, *SPAST* mutations are known to cause clinically heterogeneous manifestations, and a high interfamilial and intrafamilial variety of signs and symptoms is reported.

Although previous studies have deciphered some aspects of this variability, the genotype-phenotype correlations and some rare features of SPG4 are not fully understood. In this study, we analyzed a large cohort of SPG4 patients from Canada to better clarify the genetic and clinical spectrum of the disease.

## Methods

### Population

A total of 696 HSP patients from 431 families were recruited in eight medical centers across Canada (Montreal, Quebec, Ottawa, Toronto, Hamilton, Calgary, Edmonton, and Vancouver) as part of CanHSP, a Canadian consortium for the study of HSP. Details about the diagnosis and recruitment process has been previously reported.^10^ Clinical assessments were done and for a subset of 28 patients, the Spastic Paraplegia Rating Scale (SPRS) was measured.. For another subset of patients, brain and/or spine MRI were performed. All patients signed informed consent forms and the institutional review boards approved the study protocol.

### Genetic and data analysis

DNA was extracted from peripheral blood using a standard procedure. Initially, HSP-gene panel sequencing was performed on 379 patients. Then, 194 genetically undiagnosed patients, and additional 206 patients who were not analyzed with panel sequencing (400 patients in total from 291 families), went through whole exome sequencing (WES).

For WES, the Agilent SureSelect Human All Exon v4 kit for capture and targeted enrichment of the exome was used. To analyze the WES data, we used a list of 785 HSP-related genes or genes associated with similar neurological disorders which cause spasticity (Supplementary Table 1). Illumina HiSeq 2000/2500/4000 system was used for sequencing. Using Burrows-Wheeler Aligner (BWA), the sequence reads were then aligned against the human genome (GRCh37 assembly). We used the Genome Analysis ToolKit (GATK) and Annotate Variation (ANNOVAR) for variant calling and annotation, respectively. We excluded variant calls with a genotype quality less than 97 and less than 30x depth of coverage. Integrated Genomics Viewer was used to visually inspect the detected variants, and suspicious variants were validated by Sanger Sequencing. Sanger Sequencing was also used for assessing sporadic patients’ parents, to determine if the proband had a *de novo* mutation.

*SPAST* variants (NM_014946) were initially selected based on identifying missense and LoF alleles, including frame-shift, splice-site, nonsense, and copy number variations (CNVs) with a minor allele frequency less than 0.01 in gnomAD. The variants’ pathogenicity has been determined using VarSome, according to the American College of Medical Genetics and Genomics (ACMG) guideline. Variants classified as “Benign” and “Likely Benign”, as well as intronic splice site variants higher than ±3 were excluded from the analysis. To detect CNVs, ExomeDepth^11^ was used on WES data, followed by 48 selected samples that went through Multiplex ligation-dependent probe amplification (MLPA) testing (MRC Holland, Amsterdam, The Netherlands) to confirm or exclude suspected *SPAST* CNVs. InterPro^12^ was applied to identify domains and corresponding sites in the protein.

### Statistical analysis

To determine the association between two categorical variables, one categorical variable with one continuous variable, and two continuous variables, Pearson chi-squared test, Mann-Whitney U test, and Spearman’s rank correlation coefficient were used, respectively. *P*-values were adjusted for false discovery rate (FDR) correction as the variables were not always independent. All *P*-values were considered as significant if <0.05. SPSS was used to perform all statistical analyses.

### *In silico* structural analysis

The atomic coordinates of human spastin bound to a glutamate-rich peptide, ADP, BeF_3_ and Mg^2+^ and D. melanogaster spastin bound to a glutamate-rich peptide, ADP, ATP and Mg^2+^were downloaded from the Protein Data Bank (ID 6PEN and 6P07). The effect induced by each mutation was evaluated using the “mutagenesis” toolbox in The PyMOL Molecular Graphics System, Version 2.4.0 Schrödinger, LLC. and the DynaMut server http://biosig.unimelb.edu.au/dynamut/.

## Results

### Cohort characteristics

We identified 65 families (15.1% of all HSP families), and a total of 157 patients (22.5% of HSP patients) with SPG4. Mean AAO was 22 years (0-67, SD: 19.89), with a bimodal distribution; the first peak in the first 5 years of life, and the second peak from 35 to 44 years of age (Supplementary Figure 1). Age at onset was not associated with the severity of the disease (Spearman’s correlation coefficient; *p* = 0.934). Complex HSP was seen in 24/65 (36.9%) of the probands, and 37/157 (23.6%) of all the patients. No significant clinical differences were seen between male and females. Table 1 details the clinical presentation of the patients.

**Table 1.**
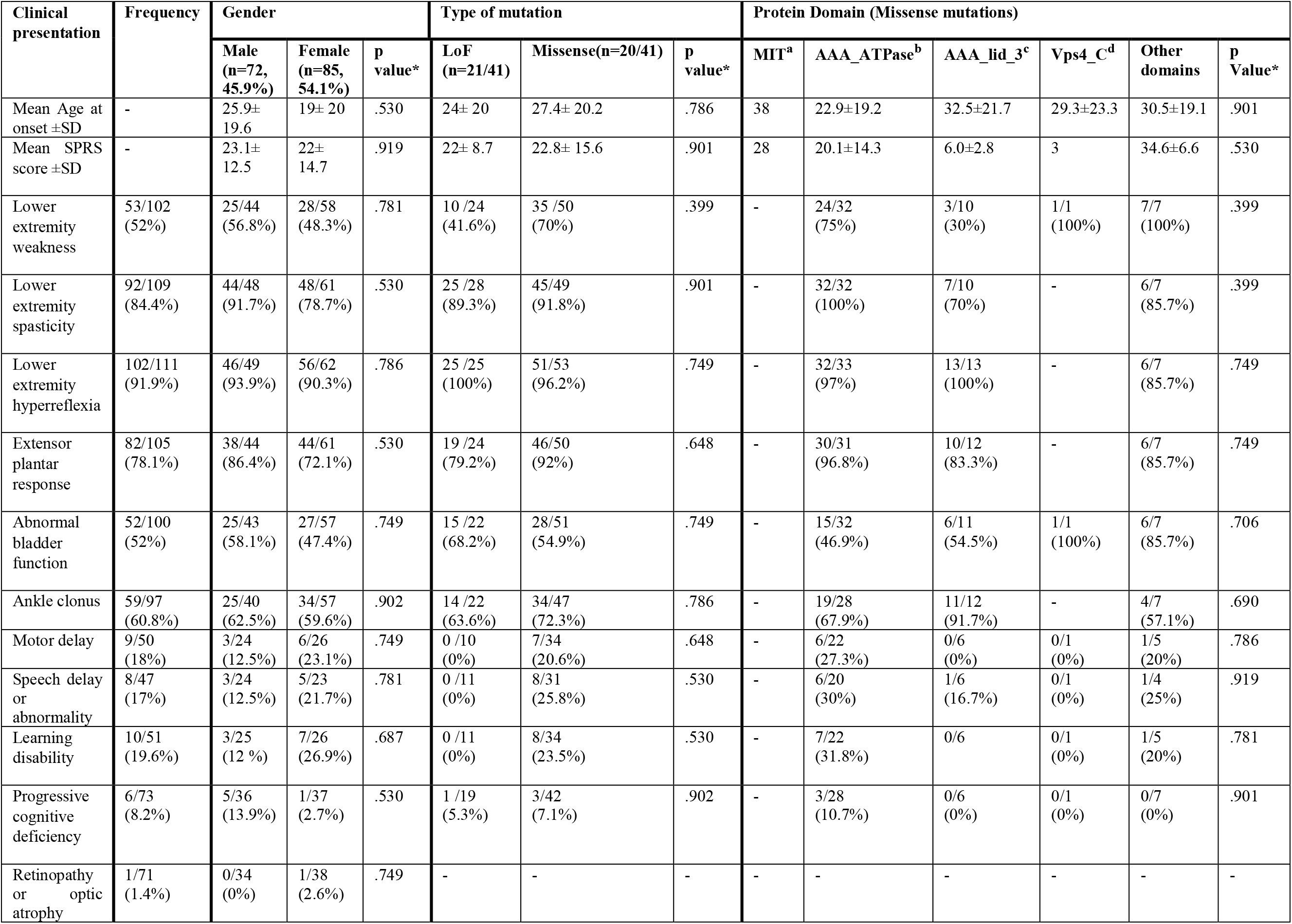
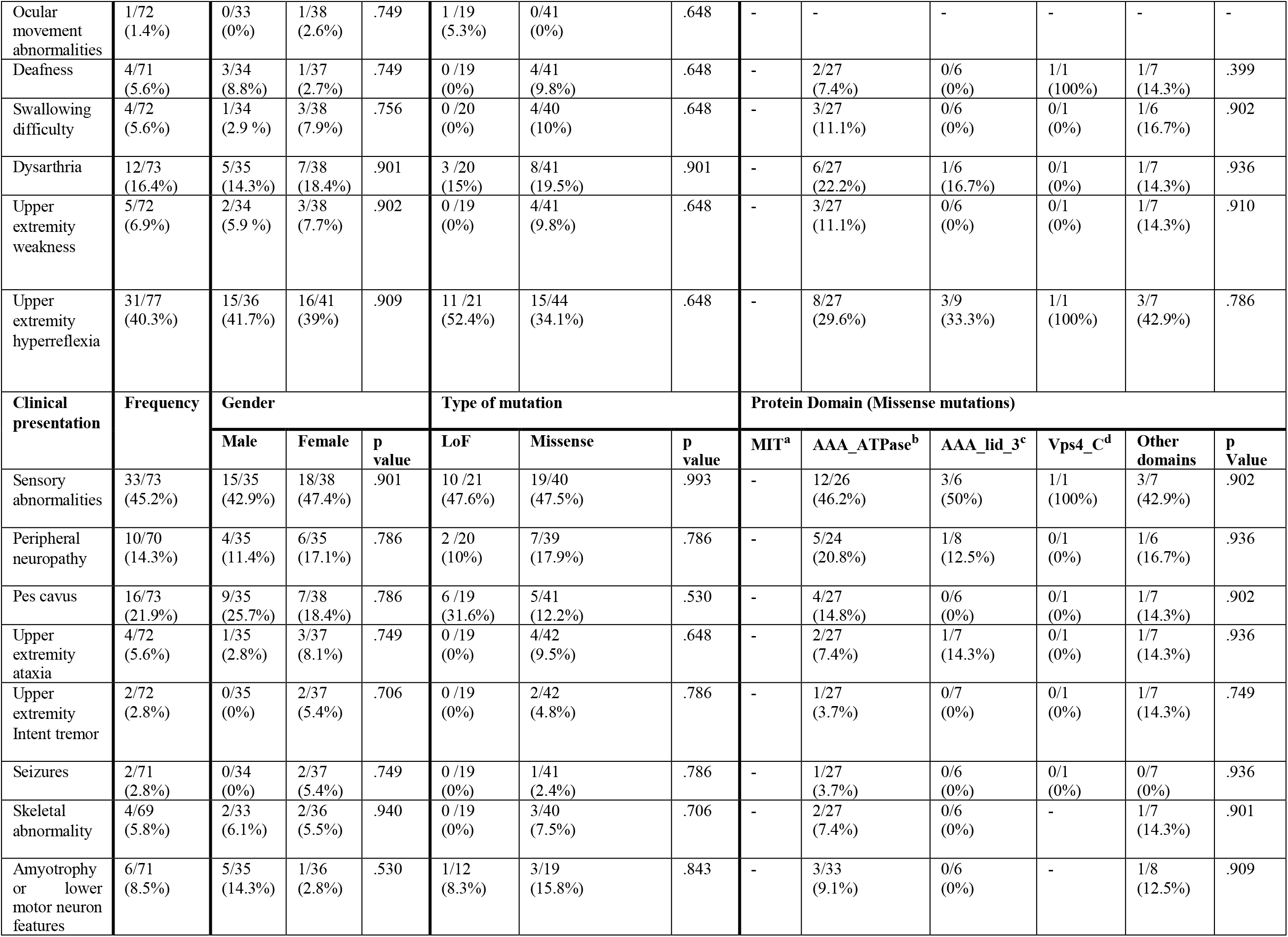
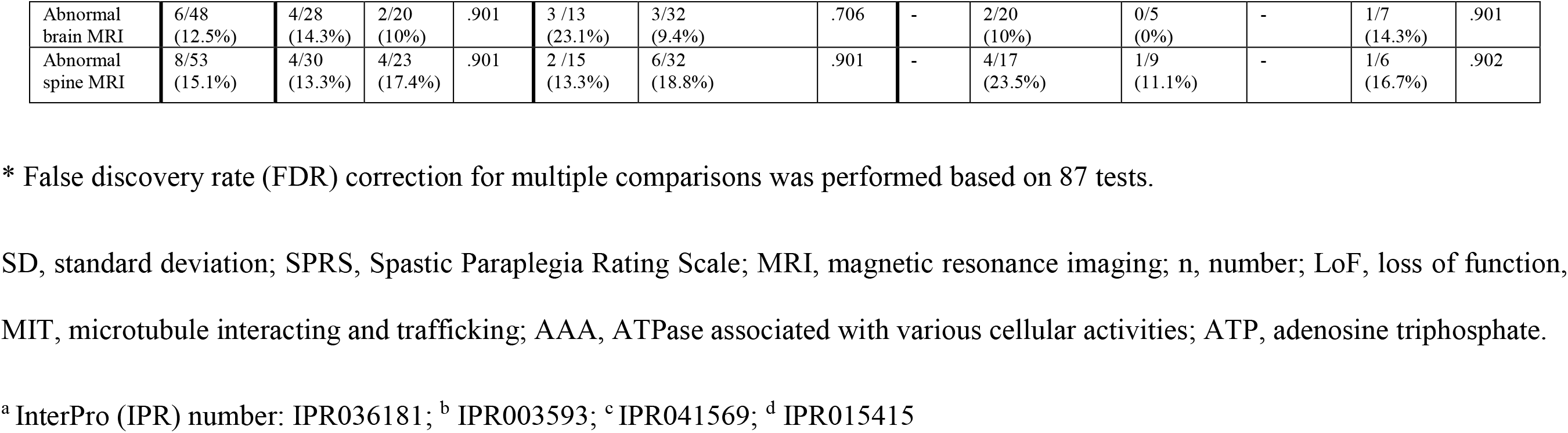
Frequency of signs and symptoms among different categories of gender, mutation type and protein domain.

We detected 41 different *SPAST* mutations in our cohort (Table 2). Most mutations (22/34, 64.7%) occurred between amino acids 374 and 567, in the AAA cassette. Among the missense mutations, 15/20 (75%) were clustered in AAA cassette, while LoF mutations were more evenly distributed across gene (Supplementary Figure 2). Presentation of the disease did not differ significantly between the two types of mutation (Table 1).

**Table 2.** Mutations identified in the current study and their characteristics.

| Nucleotide Change | Amino acid Change | Number of Families | Number of Patients | AAO (Mean) | AF in gnomAD | Pathogenicity (VarSome*) |
| --- | --- | --- | --- | --- | --- | --- |
| c.127G>T | p.(Glu43Ter) | 1 | 1 | UNK | NR | Pathogenic |
| c.153C>G | p.(Tyr51Ter) | 1 | 1 | 1 | NR | Pathogenic |
| c.231G>A | p.(Trp77Ter) | 1 | 1 | 0 | NR | Pathogenic |
| c.562delG | p.(Ala188Profs Ter8) | 1 | 1 | 38 | NR | Pathogenic |
| c.687delT | p.(Ser229Argfs Ter11) | 1 | 1 | 37 | NR | Pathogenic |
| c.869A>G | p.(Lys290Arg) | 1 | 1 | 55 | NR | Likely Pathogenic |
| c.1098delG | p.(Glu366Aspfs Ter28) | 1 | 1 | 42 | NR | Pathogenic |
| c.1111C>T | p.(Leu371Phe) | 2 | 2 | 29 | NR | Pathogenic |
| c.1112T>C | p.(Leu371Pro) | 1 | 1 | 30 | NR | Pathogenic |
| c.1139T>C | p.(Leu380Pro) | 1 | 1 | 28 | NR | Likely Pathogenic |
| c.1144_1145 insGTC | p.(Gly382_Pro383 insArg) | 1 | 2 | 15 | NR | - |
| c.1158T>G | p.(Asn386Lys) | 1 | 1 | 40 | NR | Pathogenic |
| c.1196C>T | p.(Ser399Leu) | 3 | 8 | 33 | NR | Pathogenic |
| c.1209C>A | p.(Phe403Leu) | 2 | 5 | 35 | NR | Likely Pathogenic |
| c.1212_1216del | p.(Asn405Lysfs Ter36) | 1 | 1 | 2 | NR | - |
| c.1220G>A | p.(Ser407Asn) | 1 | 1 | 1 | NR | Pathogenic |
| c.1242A>G | p.(Lys414Lys) | 2 | 3 | 9 | NR | Uncertain significance** |
| c.1276C>G | p.(Leu426Val) | 1 | 1 | 13 | NR | Pathogenic |
| c.1300C>T | p.(Gln434Ter) | 1 | 1 | 40 | NR | Pathogenic |
| c.1321+1G>A |  | 1 | 2 | UNK | NR | Pathogenic |
| c.1356_1357 insGGG | p.(Gly452dup) | 1 | 1 | 38 | NR | - |
| c.1378C>T | p.(Arg460Cys) | 1 | 1 | 40 | NR | Pathogenic |
| c.1385A>G | p.(Lys462Arg) | 1 | 1 | 14 | NR | Pathogenic |
| c.1466C>T | p.(Pro489Leu) | 1 | 4 | 19 | NR | Likely Pathogenic |
| c.1492_1493 del | p.(Arg498Alafs Ter30) | 1 | 1 | 50 | NR | Pathogenic |
| c.1493+2T>A |  | 1 | 1 | 39 | NR | Pathogenic |
| c.1495C>T | p.(Arg499Cys) | 2 | 4 | 6 | NR | Pathogenic |
| c.1496G>A | p.(Arg499His) | 4 | 4 | 2 | NR | Pathogenic |
| c.1507C>T | p.(Arg503Trp) | 3 | 4 | 34 | 4.82E-06 | Pathogenic |
| c.1537-2A>G |  | 1 | 7 | 17 | NR | Pathogenic |
| c.1610T>G | p.(Leu537Arg) | 1 | 1 | UNK | NR | Pathogenic |
| c.1676G>A | p.(Gly559Asp) | 8 | 15 | 33 | NR | Pathogenic |
| c.1684C>T | p.(Arg562Ter) | 1 | 2 | 10 | NR | Pathogenic |
| c.1685G>A | p.(Arg562Gln) | 1 | 1 | 65 | NR | Pathogenic |
| c.1729-1G>C |  | 2 | 2 | 27 | NR | Pathogenic |
| c.1741C>T | p.(Arg581Ter) | 2 | 2 | 42 | 4.81E-06 | Pathogenic |
| c.1790delG | p.(Ser597Thrfs<br>Ter3) | 1 | 3 | 4 | NR | Pathogenic |
| c.1844C>T | p.(Thr615Ile) | 1 | 1 | 30 | NR | Likely<br>Pathogenic |
| del exon 1 |  | 3 | 3 | 42 | NR | Pathogenic |
| del exons<br>5-7 |  | 1 | 2 | UNK | NR | Pathogenic |
| del exons 16-17 |  | 1 | 1 | UNK | NR | Pathogenic |

AAO, age at onset; UNK, unknown; NR, not reported; AF, allele frequency.
\* American College of Medical Genetics and Genomics (ACMG)
\*\* Mutations with uncertain significance were included with the condition of being rare and cosegregation with the disease.

### Novel *SPAST* mutations

Of the 41 different *SPAST* mutations in our cohort, six were novel, including p.(Trp77Ter), p.(Glu366AspfsTer28), p.(Gly382_Pro383insArg), p.(Phe403Leu), p.(Arg498AlafsTer30), and p.(Ser597ThrfsTer3). We performed *in silico* analysis to investigate the impact of these mutations on the structure and activity of spastin, with exception of p.(Trp77Ter), as Trp77 is not visible in any structure of spastin.

Glu366 is located in the loop_365-377_ and the frameshift mutation p.(Glu366AspfsTer28) changes all the residue subsequent to residue 365 and inserts a stop codon at position 394. This mutation truncates residues 394-616 which includes residues involved in oligomerization and ATP-binding, which will abrogate its enzymatic activity. Gly382 and Pro383 are located in the nucleotide-binding loop_382-389_.^13^ Residues Gly385 and Asn386 interact directly with the ADP, in a conformation that is shaped by the Pro383 and Pro384 (Figure 1C). The mutation p.(Gly382_Pro383insArg) lengthens this loop and likely destabilizes the interaction between the protein and the nucleotide. Phe403 forms a pi-stacking interaction with Phe439 and hydrophobic interactions with neighbouring residues (Figure 1D). Thus, mutation p.(Phe403Leu) would destabilize the domain and impair the ATPase activity.

In this structure, Arg498 and Arg499 are both involved in coordinating ADP. Arg498 is making direct interaction with the β phosphate of the nucleotide located in the active site, while Arg499 stabilizes helix_455-470_ and coordinates BeF_3_, a phosphate structural analog (Figure 1E). By homology with *D. melanogaster* spastin structure (PDB ID: 6P07), those residues seem to be also involved in ATP β and γ phosphate coordination (Figure 1F). As a result, we can hypothesize that the BeF_3_ in the active site is mimicking the γ phosphate on an ATP. Mutation p.(Arg498AlafsTer30) causes residues 498 and 499 to be serine and valine residues, which would prevent ATP stabilization. In addition, this frame shift terminates the protein after residue 528, further disrupting the ATP-binding domain. This mutation will therefore abolish the ATPase activity. Finally, mutation p.(Ser597ThrfsTer3) will induce a frame shift at residue 597 and truncate the protein at residue 600. The missing C-terminal residues form a helix involved in intersubunit interactions. This mutation would therefore destabilize the hexameric assembly, which would disrupt the ATPase activity.

### Founder French-Canadian mutations and known CNVs

The most frequent *SPAST* mutation in our cohort, p.(Gly559Asp), previously suggested to be a founder mutation in French-Canadian population^14^, was carried by 8 families (12.3%) and 15 patients in total (9.5%). Seven out of 24 French-Canadian probands (29%) harbored this mutation, while among all the patients with other or unknown ancestral backgrounds, this mutation was present in only one out of 29 probands (3.4%, *p*=0.011). Moreover, the mutation p.(Phe403Leu) was only detected in patients with French-Canadian ancestral background, in 5 patients from 2 families, suggesting it may be a founder mutation. Seven patients (4.5%) from six families (9.2%) carried a CNV (Supplementary Table 2), all of which have been previously reported. ^8^

### Earlier age at onset and specific clinical features in patients with mutations in *SPAST* p.Arg499

The range of AAO in all 4 patients from 4 different families, two of which have previously been reported^15^ with the p.(Arg499His) mutation was 1-3 years (Mann-Whitney U test; *p* = 0.003). Patients with this mutation in our cohort were more likely to present with motor delay, speech delay, dysarthria, learning disability, progressive cognitive deficits, and upper extremity weakness (Supplementary Table3).

Although statistically insignificant, cases who carried another mutation affecting the same amino acid residue, p.(Arg499Cys), also showed symptoms at a younger age (the range of AAO was 1-5 in three patients and 11-15 in one patient, *p* = 0.111). When combined together, the two mutations occurred in Arg499 locus were associated with a younger AAO (Mann-Whitney U test; *p* = 0.004). *In silico* analysis of Arg499 is discussed above, along with Arg498. Furthermore, as Arg499 stabilizes the γ phosphate, mutations p.(Arg499Cys) or p.(Arg499His) would also impede ATP binding and catalysis.

### Possible biallelic inheritance in SPG4

The *SPAST* p.(Ser44Leu) variant has been previously suggested to play modifier role in SPG4.^16, 17^ This variant was detected in one out of the 36 families that had undergone WES. From the three affected siblings, two had undergone WES, and both were heterozygous for the novel pathogenic mutation p.(Ser597ThrfsTer3), as well as for the p.(Ser44Leu) variant. The three siblings had complex HSP with disease onset in early childhood (clinical presentation of these patients is detailed in Supplementary Table 4). One unaffected parent only carried the p.(Ser44Leu) polymorphism, and the other unaffected parent was heterozygous for the pathogenic p.(Ser597ThrfsTer3) mutation and wild-type for p.(Ser44Leu) (Supplementary Figure 3). The last physical examination of the parent with the pathogenic mutation at the age between 56 to 60 revealed completely normal findings, Supporting the possible role of p.(Ser44Leu) variant as a modifier, or suggesting that extreme anticipation with undetected clinical effects in the mother could exist in this family.

Another patient with consanguineous parents, carried a homozygous mutation, *SPAST* p.(Tyr51Ter). The patient had started to show symptoms at an age range of 1-5 years and suffered from core HSP symptoms, motor and speech delay, swallowing difficulty, dysarthria, upper extremity weakness, pes cavus, and skeletal abnormalities. Her brain and total spine MRI were normal and her SPRS score was 40 when examined at an age between 16 and 20 years. The patient’s parents were both asymptomatic heterozygous carriers.

### Patients with novel or rarely reported clinical manifestations

In our cohort, four patients we report patients with deafness, ocular movement abnormality, upper extremity intention tremor, and seizure (Supplementary Table 5).

#### *De novo* cases

The parents of the three sporadic cases in our cohort did not carry the causative *SPAST* mutations. The first patient, with *SPAST* p.(Ser407Asn), had an AAO of 1-5 years. This case presented with lower extremity weakness and spasticity, upper and lower limb hyperreflexia, extensor plantar response, ankle clonus, and sensory abnormalities. They also showed signs of upper extremity weakness, motor and speech delay, dysarthria, learning disability, and swallowing difficulty. Their brain MRI was normal. The second patient had *de novo SPAST* p.(Arg499His). This patient started to show symptoms at age range of 1-5, and her symptoms included lower extremity weakness, spasticity and hyperreflexia, extensor plantar responses, and ankle clonus. Furthermore, they had motor and speech delay, dysarthria, and learning disability. They had a normal brain MRI, and their SPRS score was 37. The last *de novo* patient, also with a p.(Arg499His) mutation, also with an AAO range of 1-5 years, had an SPRS score of 28, and apart from core symptoms of HSP, had motor and speech delay, deafness, and dysarthria. The two latter patients have been previously reported.^15^

### Cases with synonymous mutation and co-occurrence of clinically isolated syndrome (CIS) and SPG4

We report three patients from two unrelated families who carried the synonymous mutation *SPAST* p.(Lys414Lys), previously reported to be pathogenic.^18^ The first patient had an AAO of between 6-10 years, and presented with lower extremity spasticity, hyperreflexia, and extensor plantar responses. The second family had two affected individuals, parent and offspring. The parent started to have difficulty walking at the age of 61-65 which led to using a cane, had marked spasticity and brisk reflexes in the lower limbs along with ankle clonus, upgoing plantar responses, urge incontinency, and mild decrease of vibration sensation in the ankles and toes. The offspring noticed difficulty walking at 21-25 years of age, which progressed slowly. This case had upper and lower limb weakness, bilateral Babinski sign and ankle clonus, and positive Hoffman sign. At an age between 31 and 35, lumbar puncture carried out due to diplopia was positive for oligoclonal bands and suggested a diagnosis of clinically isolated syndrome (CIS), which is considered as the first clinical episode in multiple sclerosis (MS).^19^ They were treated with methylprednisone, and after one month, the diplopia resolved. In addition to this patient who is a case of co-occurrence of SPG4 and CIS, from the 10 pedigrees of the families described, two demonstrate unaffected family members who have a definite or probable diagnosis of MS.

## Discussion

In this large-scale analysis of SPG4 from CanHSP, we report novel *SPAST* mutations, the possibility of a founder mutations in the French-Canadian population, novel characteristics of mutations occurring in Arg499, and potential biallelic inheritance. We also report patients with rare or novel clinical manifestations, and co-occurrence of SPG4 and CIS.

Using *in silico* analysis, we predict that the novel mutations reported in this study cause a loss-of-function of spastin, either by substantially shortening the protein, affecting its ATPase activity, or disturbing the formation of a ternary and quaternary structure necessary for catalysis. Spastin, a microtubule-cleaving enzyme involved in the cytoskeletal rearrangement, also has a role in intracellular trafficking, cytokinesis regulation and resealing of nuclear membrane. In neurons, it is involved in the regeneration of axons and axonal transport. Mutations in *SPAST* could result in a disruption of normal organelle trafficking and distribution, and thus micro-organelle accumulation in axons and swelling of axons which could result in HSP phenotype.^4, 5, 20-22^ We looked further into the three patients identified as sporadic and found out that all carried *de novo* mutations. Consistent with previous reports of more severe manifestations in patients with *de novo* mutations,^23^ all patients showed a severe form of the disease. However, it is important to note that the mutation in two of the *de novo* patients (Arg499His), which is also carried by 33% of *de novo* patients,^23^ is associated with a younger AAO. It has been suggested that amino acid 499 is located in one of the highly conserved regions of the *SPAST* gene,^24^ and mutations that occur in this region could have a significant effect on the hydrophobicity of the protein and its ability to sever microtubules.^15^

The mutation in our homozygous patient has been reported previously in a heterozygous patient;^25^ however, the presence of an additional mutation or CNVs could not have been determined in that study. Our patient with the homozygous mutation *SPAST* p.(Tyr51Ter) showed a severe form of the disease. Previously, there have been two reports of homozygous variants in *SPAST*. One, with the nonsense mutation p.(Ser545Ter), which also reports a complex and severe form of the disease,^26^ and another, with the mutation p.(Leu534Pro), who showed a pure spastic paraparesis at the age of 39.^27^ The patient from our cohort and the previously reported patient with p.(Ser545Ter) presented with a more severe form of the disease, probably because they carried nonsense mutations, while the latter patient had a missense mutation. Furthermore, in the latter study, the parents of the proband were asymptomatic carriers of the mutation.

The mode of inheritance observed in the patient with homozygous mutation and the family carrying the potential modifier mutation *SPAST* p.(Ser44Leu), may be explained by incomplete penetrance and/or anticipation rather than biallelic inheritance, which could be considered in prenatal diagnosis and genetic counselling. The synonymous mutation *SPAST* p.(Lys414Lys) we report in three patients has been previously reported to underlie the disease, and minigene assay suggested that this variant leads to an aberrant splicing effect.^18^ It is therefore important to not exclude synonymous variants when analyzing HSP specifically and other diseases in general.

The AAO distribution in our cohort followed the same trend as previously reported.^16, 25^ We did not find an association between mutation type, sex, or late onset, with the disease course.. In our data, we did not find an association between mutation type, sex, or late onset, with the disease course. Table 3 compares the frequency of symptoms not attributable to pure form of the disease in our cohort with previous studies. We report deafness in SPG4 for the first time. The co-occurrence of SPG4 and MS has been reported before; ^28, 29^ however, the results of studies assessing an association between HSP-related mutations and MS are controversial and further studies are required to verify such a relation.

**Table 3.** Frequency of symptoms not attributable to the pure form of hereditary spastic paraplegia in the present study compared to previous studies.

| **Sign/ Symptom** | **Frequency in the current study** | **Previous studies** |
| --- | --- | --- |
| Learning disabilities | 19.6% | 4.2% intellectual impairment <sup>1</sup><br>1.5% Intellectual disability <sup>2</sup> |
| Speech delay or abnormality | 17% | 4/62 (6.5%) <sup>2</sup> |
| Dysarthria | 16.4% | 3% <sup>2</sup> |
| Cognitive deficiency | 8.2% | Most frequent additional manifestation <sup>3-7</sup> |
| Upper extremity intention tremor | 2.8% | Mild arm tremor in 10% of patients <sup>2</sup> |
| Seizure | 2.8% | Case reports <sup>8-11</sup> |
| Ocular movement abnormality | 1.4% | Case reports <sup>12</sup> |
| Swallowing difficulty | 5.6% | 3% <sup>2</sup> |
| Upper limb weakness | 6.9% | Case reports <sup>5</sup> |
| Upper limb ataxia | 5.6% | Case reports <sup>5</sup> |
| Peripheral neuropathy | 14.3% | Case reports <sup>13</sup> |
| Deafness | 5.6% | - |

Our study has several limitations. For instance, in spite of being one of the largest HSP cohorts, the number of SPG4 patients was limited, especially for a genotype-phenotype correlation study. Furthermore, the families interested in participating in this study were probably those who had remained undiagnosed after the primary *SPAST* testing; therefore, the proportion of SPG4 patients in this study may be underestimated. In addition, clinical signs and symptoms for some of the patients were missing and as the study was not longitudinal, from the patients whose clinical data were collected, little follow up information was available. Also, not all comorbidities such as psychiatric disorders were assessed.

In conclusion, we report one of the largest SPG4 cohorts, with 41 different mutations including 6 novel mutations. We suggest that modes of inheritance other than autosomal dominant could be involved in SPG4. Our study sheds light on some rarely reported or unreported aspects of the disease and helps improve genetic counseling and clinical trials conducted on SPG4. Towards this goal, larger efforts and international collaborations are required.

## Data Availability

The copyright holder for this preprint is the author/funder, who has granted medRxiv a license to display the preprint in perpetuity. It is made available under a CC-BY 4.0

## Acknowledgments

We thank the patients and their families for participating in this study. We thank Patrick Dion, Daniel Rochefort, Helene Catoire, and Vessela Zaharieva for their assistance.

## Patient consent for publication

Not required

## Data availability statement

All data relevant to the study are included in the article or uploaded as online supplemental information. Anonymised data, materials and detailed protocols are available from the corresponding author on reasonable request.

## Contributors

P.V.: Research project conception, organization, and execution; statistical analysis design and execution, writing of first draft. M.A.E.: Research project conception, organization and execution; manuscript review and critique. S.A., M.F.R., G.Y., M.T., K.M.B., N.D., O.S., and E.L.: Patient recruitment and data collection. E.L, E.Y, DS, S.V., and J.F.T.: Data analysis. K.F.: Manuscript preparation. G.A.R.: Research project conception and organization; manuscript review and critique. Z.G.O.: Research project conception and organization; statistical analysis design, review and critique; manuscript review and critique.

## Supplementary Tables and Figures

**Supplementary Table 1**. Genes that are known or suspected to be involved in HSP or other similar disorders.

**Supplementary Table 2**. Copy number variations (CNVs) in the current study.

**Supplementary Table 3**. Signs and symptoms seen more frequently in patients harboring the p.(Arg499His) mutation.

**Supplementary Table 4**. Clinical manifestations of the family with probable biallelic inheritance. Patients 1 and 2 had undergone whole exome sequencing and were compound heterozygous carriers of the novel pathogenic mutation p.(Ser597ThrfsTer3), and the p.(Ser44Leu) variant. Sample from patient 3 was not available.

**Supplementary Table 5**. Characteristics of SPG4 patients with deafness.

**Supplementary Figure 1. Distribution of age at onset in the cohort**. The onset of SPG4 mostly occurs in the first five years of life, and between 35-44 years of age.

**Supplementary Figure 2. Schematic figure of the location of *SPG4* mutations in the current study**. The schematic on top represents the spastin protein. Functional domains are demonstrated in different colors, including MIT (violet), AAA-ATPase (red), AAA-lid-3 (cyan), and Ps4-C (yellow). Mutations resulting in loss of function and missense mutations are indicated with red and grey circles respectively. The number of circles in a column demonstrate the number of families that carried each mutation. The bottom schematic represents the cDNA of *SPG4*. Exons (indicated with Ex) are represented by dark blue and introns by light blue.

**Supplementary Figure 3**. Sanger sequencing results of family with modifier mutation. Mother and father carry the p.(Ser597ThrfsTer3) and p.(Ser44Leu) mutations respectively. Siblings are heterozygote for both mutations.

