## SupplementaryTable2 for "Genetic, structural and clinical analysis of spastic paraplegia 4"

**Supplementary Table 2**. Copy number variations (CNVs) in the current study.

| **CNV** | **Number of families** | **Number of patients** | **Clinical presentation** |
| --- | --- | --- | --- |
| Deletion of exon 1 | 3 | 3 | AAO:36-40  Pure HSP |
|  |  |  | AAO:41-45  HSP complicated with amyotrophy/lower motor neuron features, and peripheral neuropathy |
|  |  |  | AAO:26-30  Pure HSP |
| Deletion of exons 5-7 | 1 | 2 | AAO: 46-50  Pure HSP |
| Deletion of exons 16-17 | 1 | 1 | AAO: 36-40  Pure HSP |
| Deletion of exon 17 | 1 | 1 | AAO:36-40  Pure HSP |

AAO, age at onset; HSP, hereditary spastic paraplegia.
