## SupplementaryTable3 for "Genetic, structural and clinical analysis of spastic paraplegia 4"

| **Sign/symptom** | **Patients with Arg499His** | **Patients with other mutations** | **p value*** |
| --- | --- | --- | --- |
| Motor delay | 4/4 (100%) | 3/38 (7.9%) | 0.000018 |
| Speech delay or abnormality | 4/4 (100%) | 4/37 (10.8%) | 0.000038 |
| Learning disability | 3/4 (75%) | 5/39 (12.8%) | 0.00233 |
| Progressive cognitive deficit | 2/4 (50%) | 2/56 (3.6%) | 0.000432 |
| Upper extremity weakness | 2/4 (50%) | 2/55 (3.6%) | 0.000432 |
| Dysarthria | 4/4 (100%) | 7/56 (12.5%) | 0.000036 |

**Supplementary Table 3.** **Signs and symptoms seen more frequently in patients harboring the p.(Arg499His) mutation.**

False discovery rate (FDR) corrected.
