## SupplementaryTable4 for "Genetic, structural and clinical analysis of spastic paraplegia 4"

**Supplementary Table 4.** **Clinical manifestations of the family with probable biallelic inheritance**. Patients 1 and 2 had undergone whole exome sequencing and were compound heterozygous carriers of the novel pathogenic mutation *SPAST* p.(Ser597ThrfsTer3), and the *SPAST* p.(Ser44Leu) variant. Sample from patient 3 was not available.

| **Pt** | **AAO** | **Spasticity** | **Delayed Motor**  **Milestones** | **Speech**  **Delay** | **Dysarthria** | **Learning**  **difficulty** | **Dysphagia** | **UE weakness** | **LE**  **Weakness** | **HR** | **Hoffman**  **(R/L)** | **Babinski sign**  **(R/L)** |
| --- | --- | --- | --- | --- | --- | --- | --- | --- | --- | --- | --- | --- |
| 1 | 1-5 | +  (LE>>UE) | - | - | - | - | - | + | + | + (LE>UE) | -/- | +/+ |
| 2 | Early childhood | + | + | + | Mild | - | - | - | + | + (LE>UE) | +/+ | +/+ |
| 3 | Early  childhood | + (LE>>UE) | + | + | - | + | Mild | + | + | + (LE>UE) | +/+ | +/+ |

Pt, patient; AAO, age at onset; LE, lower extremity; UE, upper extremity; HR, hyperreflexia; R, right; L, left.
