## SupplementaryTable5 for "Genetic, structural and clinical analysis of spastic paraplegia 4"

**Supplementary Table 5.** **Charactristics of SPG4 patients with novel or rarely reported clinical manifestations.**

| **Novel/rare manifestation** | **Details** | **Mutation** | **Age at onset** | **SPRS**  **score** | **Other signs/symptoms attributed to complex HSP** | **Pedigree*** |
| --- | --- | --- | --- | --- | --- | --- |
| Deafness | - | p.Asn386Lys | 41-45 | NA | - | C |
|  |  | p.Lys290Arg | 51-55 | 42 | - | D |
|  |  | p.Arg499His | 1-5 | 28 | Motor delay, speech delay, dysarthria | - (*De novo*) |
|  |  | p.Arg581Ter | 31-35 | 3 | - | Not available |
| Ocular movement abnormality  (Could indicate cerebellar involvement) | Extraocular movement showed hypometric saccades, pursuit smooth,  Mild horizontal gaze evoked nystagmus | p.(Gln434Ter) | NA | NA | - | E |
| Upper extremity intention tremor  (Could indicate cerebellar involvement) | - | p.(Arg503Trp) | 46-50 | 34 | Swallowing difficulty, peripheral neuropathy, upper extremity ataxia | F |
|  |  | p.(Leu371Pro) | 26-30 | 39 | Upper extremity ataxia | NA |
| Seizure | Generalized seizure starting at age range 41-45,  Two normal cranial CT scans at age range 66-70 | p.(Pro489Leu) | 21-25 | NA | - | G |
|  | Episodes of atypical seizures at age range 56-60 | c.1321+1G>A | 1-5 | NA | - | H |

NA, not available; CT, computed tomography.

*Supplementary Figure 4.
