## Supplementary figures and images for "Genetic, structural and clinical analysis of spastic paraplegia 4"

### SupplementaryFigure1

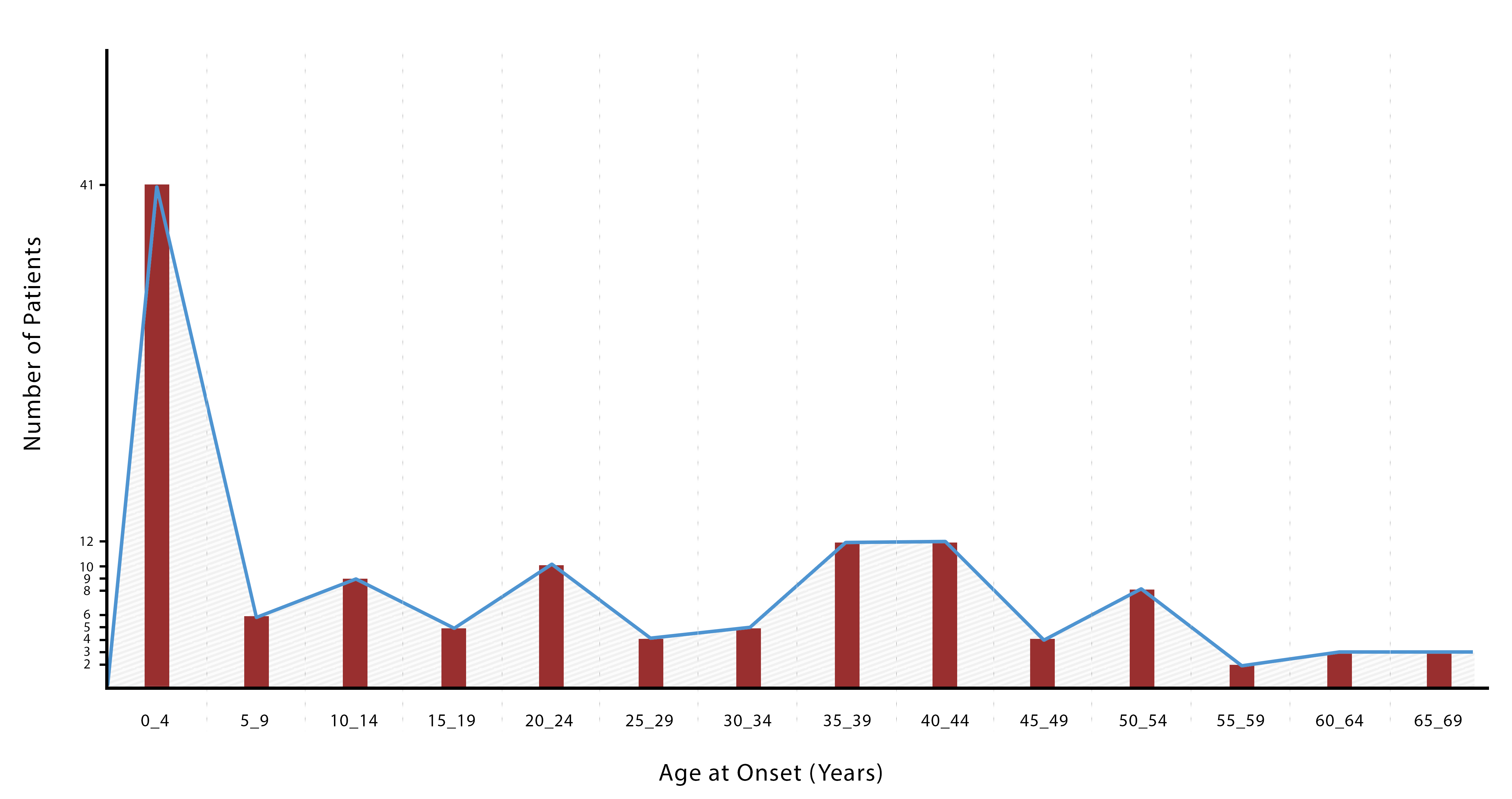

### SupplementaryFigure2

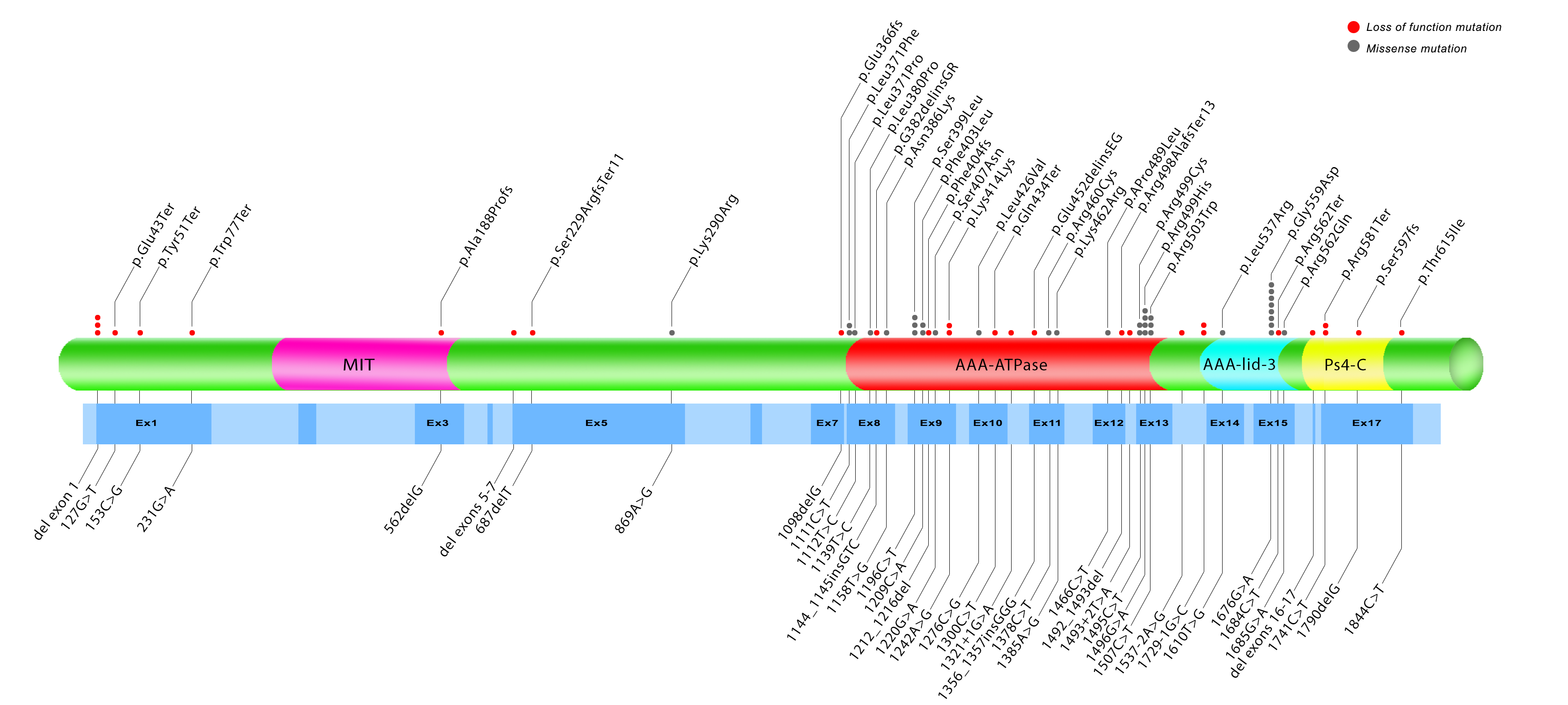

### SupplementaryFigure3

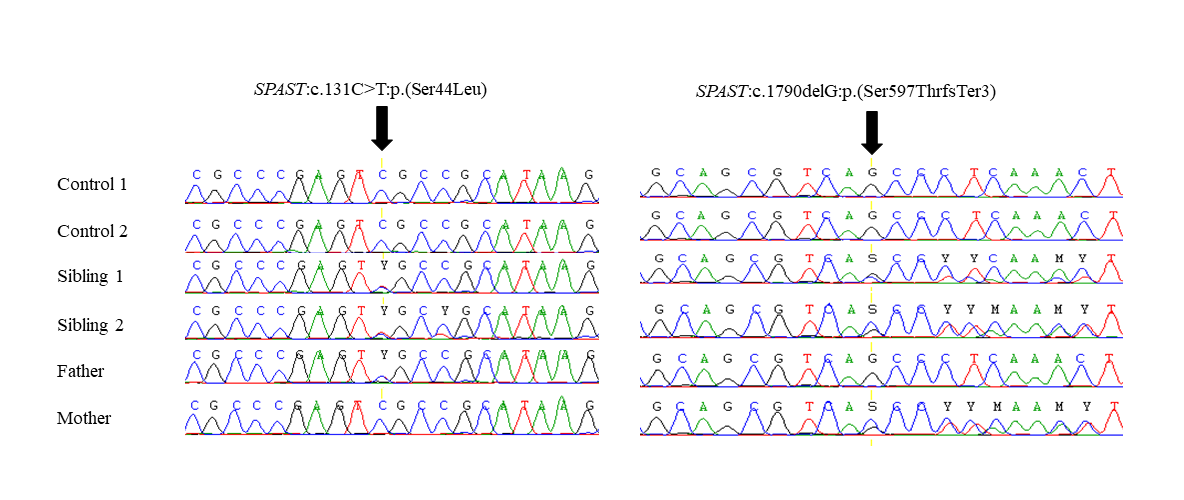
